# Evaluation of six commercial SARS-CoV-2 Enzyme-Linked Immunosorbent assays for clinical testing and serosurveillance

**DOI:** 10.1101/2021.01.21.21250249

**Authors:** Suellen Nicholson, Theo Karapanagiotidis, Arseniy Khvorov, Celia Douros, Francesca Mordant, Katherine Bond, Julian Druce, Deborah A Williamson, Damian Purcell, Sharon R Lewin, Sheena Sullivan, Kanta Subbarao, Mike Catton

## Abstract

**Background:** Serological testing for SARS-CoV-2 complements nucleic acid tests for patient diagnosis and enables monitoring of population susceptibility to inform the COVID-19 pandemic response. As we move into the era of vaccines, the detection of neutralising antibody will become increasingly important. Many serological tests have been developed under emergency use authorization, but their reliability remains unclear.

**Methods:** We evaluated the performance of six commercially-available Enzyme-linked Immunosorbent Assays (ELISAs), including a surrogate virus neutralization test, for detection of SARS-CoV-2 immunoglobulins (IgA, IgM, IgG), total or neutralising antibodies and a subset of results were compared to microneutralisation.

**Results:** For sera collected > 14 days post-symptom onset the Wantai total Ab performed best with highest sensitivity 100% (95% confidence interval: 94.6-100) followed by 93.1% for Euroimmun NCP-IgG,93.1% for GenScript Surrogate Virus Neutralization Test, 90.3% for Euroimmun S1-IgG, 88.9% for Euroimmun S1-IgA and 83.3% for Wantai IgM. Specificity for the best performing assay was 99.5% and for the lowest 97.1%.

**Conclusion:** Wantai ELISA, detecting total immunoglobulins against SARS-CoV-2 receptor binding domain, had the best performance. Antibody target, timing and longevity of the immune response, and the objectives of testing should be considered in test choice. ELISAs should be used within a confirmatory testing algorithm to ensure reliable results. ELISAs provide high quality results, with flexibility for test numbers without the need for manufacturer specific analyzers.

## INTRODUCTION

Severe Acute Respiratory Syndrome Coronavirus 2 (SARS-CoV-2) the causative agent of Coronavirus disease 2019 (COVID-19) has infected > 71 million people and caused > 1.6 million deaths globally as of 18 December 2020 [1]. COVID-19 manifests as an acute respiratory illness, although asymptomatic infections occur [2].

Laboratory diagnosis of COVID-19 primarily relies on detection of viral RNA by RT-PCR in respiratory tract samples or antigen tests, the sensitivity of which declines as infection resolves. In contrast, antibody (Ab) is detected in most individuals 10-15 days following onset of symptoms [3]. Serological testing can detect previous infection in people who have recovered without RT-PCR testing, who are RT-PCR negative [4, 5], or whose RT-PCR results are difficult to interpret [6]. Serology may also inform our understanding of antibody longevity and quantification of neutralising antibody (nAb) response in vaccine trials [6, 7, 8]. As the pandemic progresses estimates of prior exposure and population prevalence from serological assays will play an increasingly important role for public health decision making [9, 10]. However, there is substantial variation in assay performance and correlation to neutralising antibodies [11].

In this study we evaluate the sensitivity and specificity of six commercially available serological ELISA tests using either spike (S), nucleocapsid (NCP) or receptor binding domain (RBD) antigens for detection of specific isotypes of SARS-CoV-2 (IgA, IgM, IgG), total Ab or nAb (using a surrogate virus neutralization test (sVNT)). We also compared a subset of ELISA results to microneutralisation (MN), the current gold standard assay for SARS-CoV-2-specific antibody.

## METHODS

### Study Design

A retrospective study evaluating the sensitivity and specificity of commercially available ELISAs for detection of Abs to SARS-CoV-2 virus on well pedigreed sera.

### Evaluation serum samples

Sera from three patient groups were assembled to assess aspects of assay performance: (1) Infected group: sera from patients with prior RT-PCR confirmed SARS-CoV-2 infection for the assessment of sensitivity. Specificity was calculated for (2) Population group: pre-pandemic sera representing the Victorian population, collected between 2011 and 2018 and (3) Cross-reactive group: pre-pandemic sera for assessment of potential cross-reactivity; from patients with seasonal coronavirus, SARS-CoV1, MERS-CoV or other non-COVID-19 acute infections (see Table 1 and Table 3 for the full list).

**Table 1.**
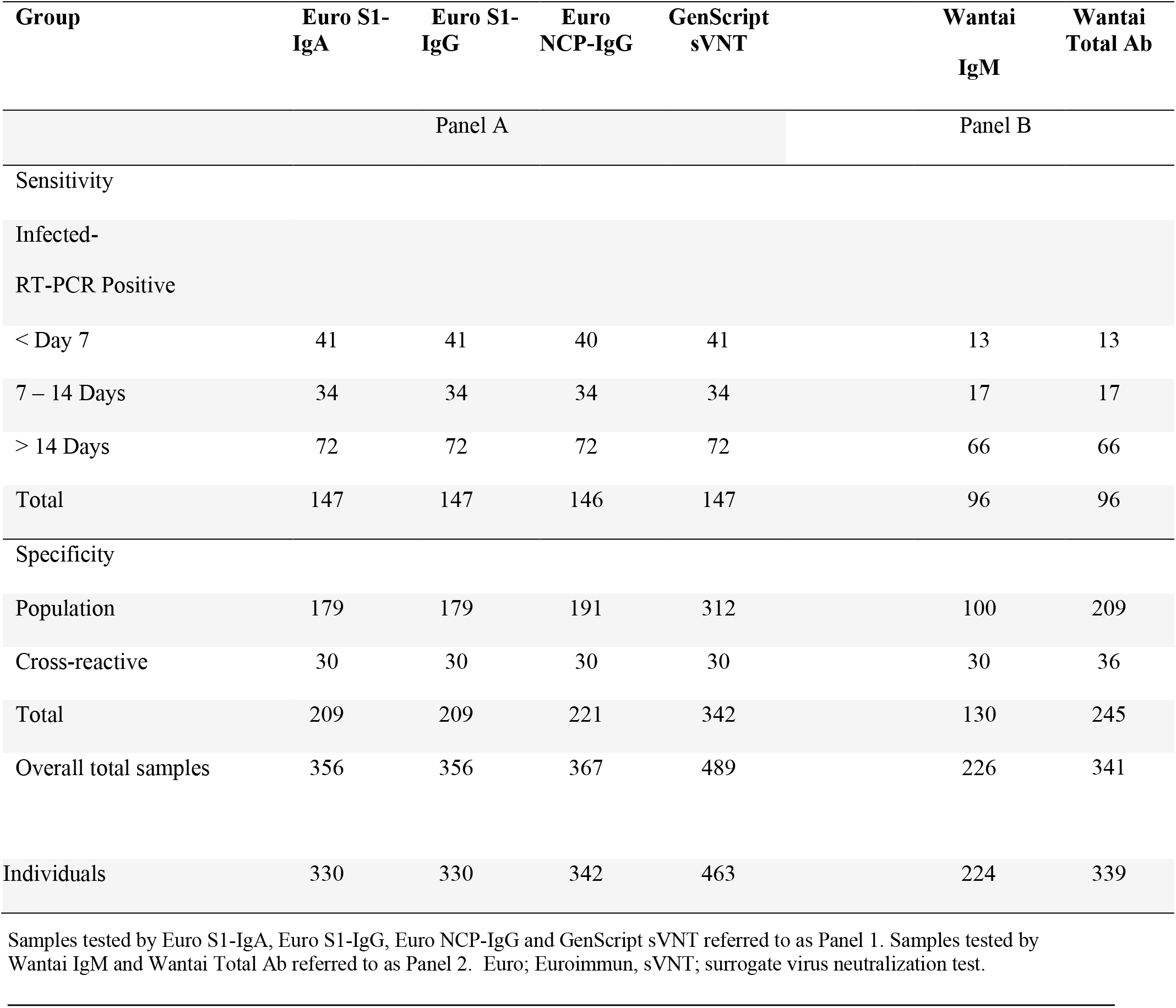
Description of Evaluation Samples for the assays

Samples were obtained from the Victorian Infectious Diseases Reference Laboratory (VIDRL) at The Peter Doherty Institute for Infection and Immunity and Royal Melbourne Hospital (RMH), Victoria, Australia (Table 1).

### Testing protocols

#### RT-PCR

SARS-CoV-2 RNA was detected in respiratory swabs by RT-PCR and results provided by both institutions using either in-house RT-PCR with previously published primers [12] used by VIDRL, or a commercial Coronavirus Typing assay, used by RMH [13]. All positive samples were confirmed by a second assay/target.

#### ELISA

ELISA testing was performed in the Serology Laboratory at VIDRL following manufacturers’ Instructions for Use (IFU) with results reported semi-quantitatively as either a signal/cut-off ratio (Euroimmun and Wantai) or percentage inhibition (sVNT).

After initial sVNT testing we also repeated testing if results were within 10% of the IFU-suggested cut-off (18%-22%). Assay performance characteristics were assessed using the panels described previously. Testing was performed manually though high throughput testing could be performed using robotic platforms. Intra-assay variability was calculated by testing 10 in-house quality control sample replicates within the same microtitre plate and inter-assay variability by testing on different plates on different days using the same kit lot number; results are reported as a coefficient of variation (CV).

### Euroimmun anti-SARS-CoV-2 S1-IgG, S1-IgA and anti-SARS-CoV-2 NCP-IgG

(Euroimmun Medizinische Labordiagnostika, Lubeck, Germany)

These are indirect ELISAs for detection of immunoglobulin (Ig) class IgG or IgA against SARS-CoV2 antigens. Wells are coated with either (i) S protein S1-domain or (ii) modified NCP. SARS-CoV-2 binding antibodies are detected using enzyme-labelled anti-human-IgG or anti-human-IgA conjugates and a colourmetric substrate and are read spectrophotometrically.

### Wantai SARS-CoV-2 total Ab

(Beijing Wantai Biological Pharmacy Enterprise, Beijing China)

This is a two-step incubation antigen ‘sandwich’ assay detecting total antibodies binding the SARSCoV-2 RBD within the S1-subunit of S protein. Patient antibody to SARS-CoV-2 that binds antigen coated on the plate, is bound by horseradish peroxidase (HRP) antigen conjugate forming an antigen-antibody-antigen-HRP complex detectable by colourmetric substrate and read spectrophotometrically.

### Wantai SARS-CoV-2 IgM

This is a capture ELISA for detection of IgM-class antibodies to SARS-CoV-2 virus. Anti-µ chain antibodies on the plate capture patient IgM antibodies; detection is by recombinant SARS-CoV-2antigen-HRP-conjugate followed by a colourmetric substrate read spectrophotometrically.

### GenScript SARS-CoV-2 Surrogate Virus Neutralization Test (sVNT)

(GenScript USA, Inc. New Jersey, USA)

This is a species and isotype independent blocking ELISA which mimics virus neutralisation, detecting circulating neutralising SARS-CoV-2 antibodies that block the interaction between the viral spike RBD and host angiotensin converting enzyme 2 (ACE2) cell surface receptor. HRP-conjugated recombinant SARS-CoV-2 RBD fragment binds to any circulating nAb to RBD preventing capture by the hACE2 protein on the well which is subsequently removed in the following wash step. Substrate reaction incubation time is determined by temperature; ideal reaction temperature and time in the IFU are 25°C for 15 minutes. For temperatures lower than 25°C, the time can be extended. At 15 minutes our control values did not meet the assay validity criteria, but at 20 minutes they fell within the acceptable ranges. Colour intensity is inversely dependent on the titre of anti-SARS-CoV-2 nAbs. We report three types of alternative estimates: (i) 20% cut-off without retesting (as per the IFU), (ii) 20% cut-off with retesting for equivocal results (18-22%) and (iii) 25% cut-off without retesting.

### Microneutralisation assay

In a subset of sera, a comparison was performed using an in-house MN assay and the commercial ELISAs. To compare MN sensitivity and specificity to the Euroimmun and sVNT assays, ninety samples from RT-PCR positive patients were assessed (Panel A). Seventy samples were assessed for comparison of MN to the Wantai assays, which require a higher volume of sera than the other assays (Panel B).

The MN assay was performed using an in-house assay as described previously [13]. Briefly, the ability of serial 2-fold dilutions of sera to neutralize the infectivity of 100 median tissue culture infectious doses of SARS-CoV-2 was assessed by inhibition of viral cytopathic effect in Vero cells.

The nAb titre was calculated using the Reed/Muench method [14, 15].

### Statistical analysis

All statistical analyses were conducted using R version 4.0.2 [16]. Responses were assessed in each of the 3 patient groups (Infected, population, cross-reactive). Sensitivity was estimated using the infected group separately for each of three categories from time of symptom onset: (<7 days, 7-14 days, >14 days). Each observation was treated as independent within each subgroup. Specificity was estimated separately for the population group and the cross-reactive group. The 95% confidence intervals were generated using the exact binomial Clopper-Pearson method with PropCIs R package [17]. Sensitivity estimates (point estimates and CI bounds) were averaged across the onset categories to give the “averaged” sensitivity estimate. Point estimates and interval bounds for the sensitivity and the population group specificity were used to calculate the corresponding estimates and bounds for PPV and NPV at different theoretical levels of population prevalence (0.1%, 0.5%, 1%, 5%, 10% and 20%).

### Ethics

Project ethical approval for RMH specimens was obtained from Melbourne Health Human Research Ethics Committee (RMH HREC QA2020052). The in-house panel consisted of anonymised excess diagnostic specimens sent to VIDRL for COVID-19 testing and the ‘VIDRL Serum Reference Collection’.

## RESULTS

### Sensitivities and specificities of six ELISA assays

Performance characteristics of the six commercial assays are shown in (Table 2). Increased sensitivity as well as increased antibody level, reported as index values or % inhibition, was observed across all timeframes for all assays (Figure 2). For comparative assay sensitivity and specificity see (Figure 1).

**Table 2.** Performance Characteristics of assays with RT-PCR, all time points and average.

| Group | Euro<br>S1 IgA | Euro<br>S1 IgG | Euro<br>NCP IgG | GenScript<br>sVNT<br>(20% c/o)^<br>(20% c/o/equ)*<br>(25% c/o)# | Wantai<br>IgM | Wantai<br>Total Ab |
| --- | --- | --- | --- | --- | --- | --- |
| Sensitivity %<br>[95% CI] | Panel A |  |  |  | Panel B |  |
| Infected-RT-PCR Pos |  |  |  |  |  |  |
| Averaged | 58.0 [44.4-71.1] | 46.7 [35.4-59.6] | 51.4 [39.4-64.2] | 54.4 [42.3-66.7]^*# | 61.6 [41.8-80.8] | 71.6 [52.8-87.2] |
| < Day 7 | 29.3 [16.1-45.5] | 14.6 [5.6-29.2] | 20.0 [9.1-35.6] | 17.1 [7.2-32.1]^*# | 30.8 [9.1-61.4] | 38.5 [13.9-68.4] |
| 7 – 14 Days | 55.9 [37.9-72.8] | 35.3 [19.7-53.5] | 41.2 [24.6-59.3] | 52.9 [35.1-70.2]^*# | 70.6 [44-89.7] | 76.5 [50.1-93.2] |
| > Day 14 | 88.9 [79.3-95.1] | 90.3 [81.0-96.0] | 93.1 [84.5-97.7] | 93.1 [84.5-97.7]^*# | 83.3 [72.1-91.4] | 100 [94.6-100] |
| Specificity %<br>[95% CI] |  |  |  |  |  |  |
| Population | 95 [90.7-97.7] | 97.2 [93.6-99.1] | 99 [96.3-99.9] | 97.1 [94.6-98.7]^<br>99.4 [97.7-99.9]*<br>100 [98.8-100]# | 99.0 [94.6-100] | 99.5 [97.4-100] |
| Cross-reactive<br>assessment | 86.7 [69.3-96.2] | 90.0 [73.5-97.9] | 83.3 [65.3-94.4] | 96.7 [82.8-99.9]^*# | 96.7 [82.8-99.9] | 97.2 [85.5-99.9] |
| PPV % (Best-worst)<br>0.5% prevalence<br>>Day14 | 8.2 [4.1-17.0] | 14.0 [6.0-34.6] | 30.9 [10.2-79.4] | 13.8 [7.1-26.8]^<br>42.2 [15.6-86.3]*<br>100 [25.7-100]# | 29.5 [6.2-94.8] | 51.2 [15.3-97.6] |
| NPV % (Best-worst)<br>0.5% prevalence<br>>Day14 | 99.9 [99.8-100] | 99.9 [99.9-100] | 100 [99.9-100] | 100 [99.9- 100]^*# | 99.9 [99.9-100] | 100 [100-100] |
| PPV (Best-worst)<br>10% prevalence<br>>Day 14 | 66.3 [48.6-82.0] | 78.2 [58.4-92.1] | 90.8 [71.6-98.8] | 77.9 [63.0-89.0]^<br>94.2 [80.4-99.3]*<br>100 {88.4-100}# | 90.3 [59.5-99.8] | 95.9 [79.9-99.1] |
| NPV (Best-worst)<br>10% prevalence<br>>Day 14 | 98.7 [97.5-99.4] | 98.9 [97.8-99.6] | 99.2 [98.2-99.7] | 97.8 [94.1-99.7]^*# | 98.2 [96.8-99.1] | 100 [99.4-100] |
| For all calculations, equivocal results were treated as positive. 95% CI, 95% confidence interval; Best-worst; best and worst- case intervals – intervals obtained by setting the corresponding sensitivity and specificity estimates to their upper/lower interval bound) Euro; Euroimmun. Averaged; average sensitivity across the symptom onset categories.PPV; Positive Predictive Value. NPV; Negative Predictive Value, c/o; cut-off, ^ [20% c/o; s-vnt ≥ 20% cut-off, no repeats], *[20% c/o/equiv; s-vnt ≥ 20% cut-off + repeats for equivocals (18%-22%)], # [25% c/o; ≥ 25% cut-off, no repeats]. |  |  |  |  |  |  |

**Table 3.** Cross-reactivity results for non-COVID-19 mixed infection pre-pandemic group

|  | Euro S1-IgA | Euro S1-IgG | Euro NCP-IgG | GenScript sVNT | Wantai IgM | Wantai Total Ab |
| --- | --- | --- | --- | --- | --- | --- |
| **Antibodies against** |  |  |  |  |  |  |
| SARS-CoV-1 | 1/1 | 1/1 | 1/1 | 1/1 | 1/1 | 1/1 |
| MERS-CoV | 0/9 | 0/9 | 1/9 | 0/9 | 0/9 | 0/9 |
| Seasonal hCoV | 0/5 | 0/5 | 0/5 | 0/4 | 0/2 | 0/2 |
| Mycoplasma | 1/4 | 1/4 | 2/4 | 0/4 | 0/2 | 0/2 |
| Parvovirus, Parvo/HCV | 0/4 | 0/4 | 0/4 | 0/4 | 0/5 | 0/2 |
| CMV | 1/2 | 1/2 | 1/2 | 0/2 | 0/7 | 0/5 |
| Dengue | 1/5 | 0/5 | 0/5 | 0/5 | 0/6 | 0/7 |
| EBV | 0 | 0 | 0 | 0 | 0/4 | 0/4 |
| Flu A, Adenovirus | 0 | 0 | 0 | 0 | 0/1 | 0/1 |
| Hep A, B, C, Syphilis, HIV | 0 | 0 | 0 | 0 | 0/6 | 0/6 |
| **Total** | **4/30** | **3/30** | **5/30** | **1/30** | **1/30** | **1/36** |
| **% Positive** | **13.3%** | **10.0%** | **16.7%** | **3.3%** | **3.3%** | **2.8%** |

Euro; Euroimmun; SARS-CoV-1; Severe Acute Respiratory Syndrome Coronavirus 1; MERS-CoV; Middle East respiratory syndrome, CMV; cytomegalovirus, Parvo; parvovirus, HCV; Hepatitis C virus, EBV; Epstein Barr virus, Flu A; Influenza A virus, Hep; hepatitis, HIV; human immunodeficiency virus.

**Figure 1.**
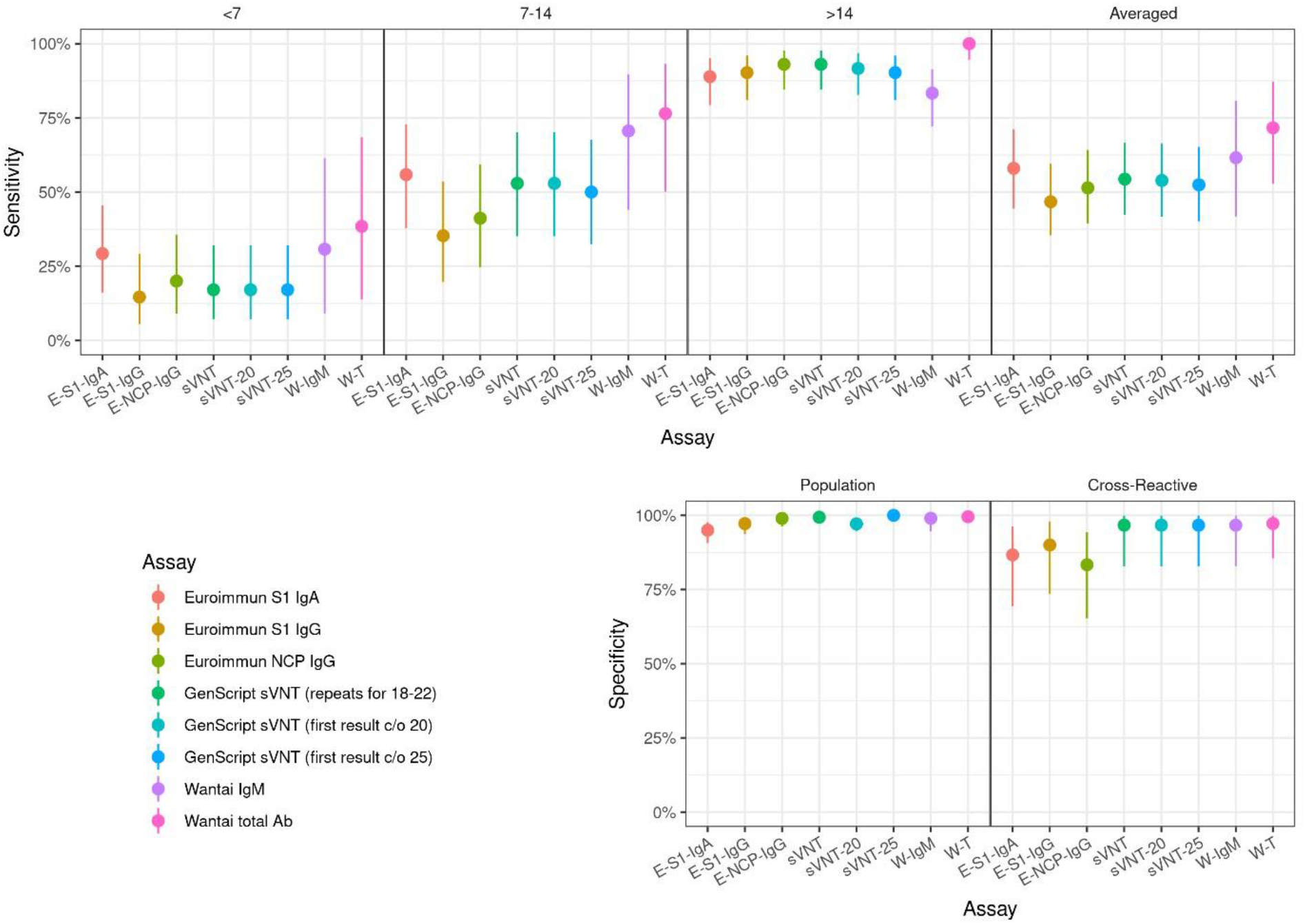
Comparative analysis of assay sensitivity post symptom onset < 7 days, 7-14 days, ≥ 14 days and averaged. Comparative analysis of assay specificity in cross-reactive assessment and population groups Abbreviations: E-S1-IgA, Euroimmun S1-IgA; E-S1-IgG, Euroimmun S1-IgG; E-NCP-IgG, Euroimmun NCP-IgG; sVNT, GenScript surrogate virus neutralization test 20% cut-off with repeat testing for equivocal results (18-22); sVNT-20 with 20% cut-off with no repeat testing; sVNT-25 with 25% inhibition cut-off and no repeat testing; W-IgM, Wantai IgM, W-T, Wantai Total Ab.

**Figure 2.**
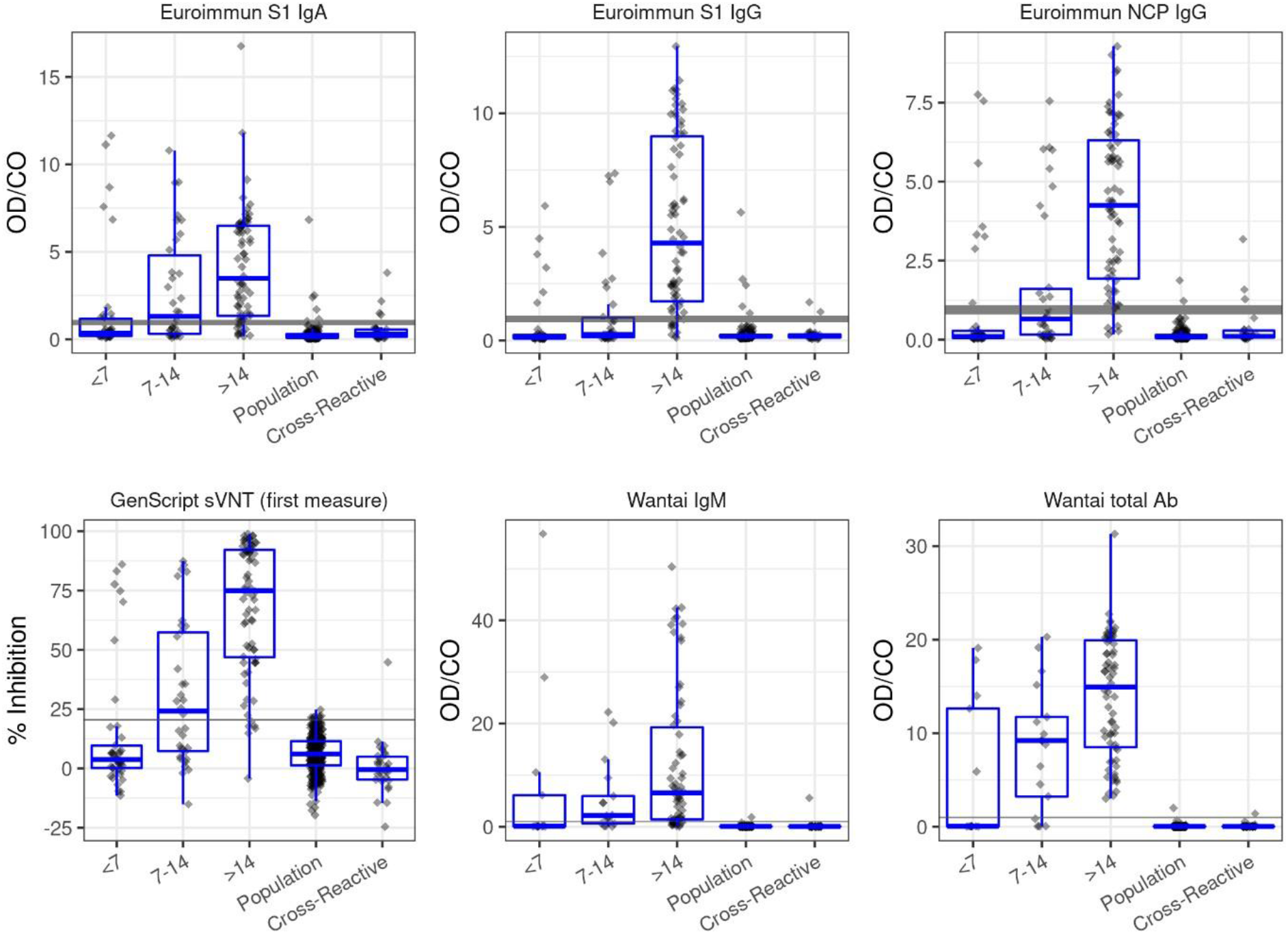
Boxplots of data distribution, as signal/cut-off value for each assay for the RT-PCR Positive and control sera by days post symptom onset; <7 days, 7-14 days and >14 days. Boxes represent median values and interquartile range, and whiskers represent largest and smallest values. Grey horizontal lines represent the cut-off value, and the shaded grey indicates the equivocal/borderline zone. Measure is index value (sample optical density (OD) value/ cut-off OD) for all assays except sVNT. sVNT measure is %inhibition, calculated as per IFU.

#### Euroimmun S1 IgA, S1 IgG and NCP IgG

Sensitivity was assessed using sera from subjects with RT-PCR proven COVID-19 disease (Table 2).

When sera collected > 14 days post-symptom onset were considered, sensitivity was 88.9% (79.395.1) for S1-IgA, 90.3% (81.0-96.0) for S1-IgG and 93.1% (84.5-97.7) for NCP-IgG. The averaged sensitivity across all timeframes was 58.0% (44.4-71.1) for S1-IgA, 46.7% (35.4-59.6) for S1-IgG and 51.4% (39.4-64.2) for NCP IgG.

Specificity was assessed using the population and cross-reactive groups (Table 2). Specificity when testing the pre-pandemic population group was lowest at 95.0% (90.7-97.7) using the S1-IgA, 97.2% (93.6-99.1) using S1-IgG and highest 99.0% (96.3-99.9) using NCP-IgG. In the cross-reactive assessment group, the lowest specificity was 83.3% (65.3-94.4) using NCP-IgG, 86.7% (69.3-96.2) using S1-IgA and highest using S1-IgG at 90% (73.5-97.9). Initial testing of the S1-IgA kit gave poorer specificity than reported here; however, with the introduction of a new buffer by Euroimmun, retrospective testing of pre-pandemic samples showed an 8.5% specificity improvement.

Cross-reactivity was observed with sera containing anti-SARS-CoV-1 antibody (all tests), anti-MERS antibody (only NCP IgG, 1/9 samples), and sera positive for mycoplasma and CMV (all tests) and dengue (only S1-IgA, 1/5 samples) antibodies. No cross-reactivity was observed to sera positive for parvovirus-B19 and seasonal coronavirus antibodies (Table 3).

#### GenScript SARS-CoV-2 sVNT

For the COVID-19 RT-PCR positive group, sensitivity of the sVNT was 93.1% (84.5-97.7) when sera collected > 14 days from symptom onset were considered and was 54.4% (42.3-66.7) across all time periods (Table 2). The assay specificity was 97.1% (94.6-98.7) when the pre-pandemic population group were tested. All other assays required re-testing of samples with results in the ‘equivocal zone’, 10-20% of the assay cut-off, depending on the assay. Because the sVNT had similar intra-assay and inter-assay variability to the other assays, we wanted to see if specificity improved with retesting of samples in the 10% equivocal zone. When the pre-pandemic population group sera were tested with repeat testing of equivocal zones specificity, increased to 99.4% (97.7-99.9). With a 20% cut-off and equivocal zone repeat testing 2/312 samples gave a false positive result (both MN negative), both were close to the 20% inhibition cut-off (22.5%, 21%). Because the IFU suggests setting a population specific cut-off, we considered a range of thresholds (Supplementary Figure 1a, 1b, 1c & Table 1a, 1b). Using a cut-off value of 25% increased specificity and PPV without changing the sensitivity and NPV (Table 2). In the cross-reactivity assessment group cross-reactivity was observed with sera containing antibody to SARS-CoV-1 (Table 3).

#### Wantai Total Ab and IgM

The averaged sensitivity for the Wantai Total Ab was 71.6% (52.8-87.2) and for the IgM assay 61.6% (41.8-80.8) which improved to 100.0% (94.6-100.0) and 83.3% (72.1-91.4), respectively, when restricted to sera collected > 14 days post-symptom onset. (Table 2).

The specificity for the Wantai Total Ab was 99.5% (97.4-100.0) and for IgM was 99.0% (94.6-100.0) in the population group and for the cross-reactive group these were 97.2% (85.8-99.9) and 96.7% (82.8-99.9), respectively. Cross-reactivity was observed with sera containing antibody to SARS-CoV1 (Table 3).

### Positive Predictive Value (PPV) and Negative Predictive value (NPV)

PPV and NPV were calculated across a range of population prevalence estimates (Figure 3). In a low prevalence setting like ours, assuming population prevalence of 0.32% for Victoria and 0.11% for Australia [18] PPV for the best performing assay, Wantai Total Ab was 40.2% and 18.7%, respectively while the NPV was 100% for sera collected >14 days post-symptom onset (Supplementary Table 2).

**Figure 3.**
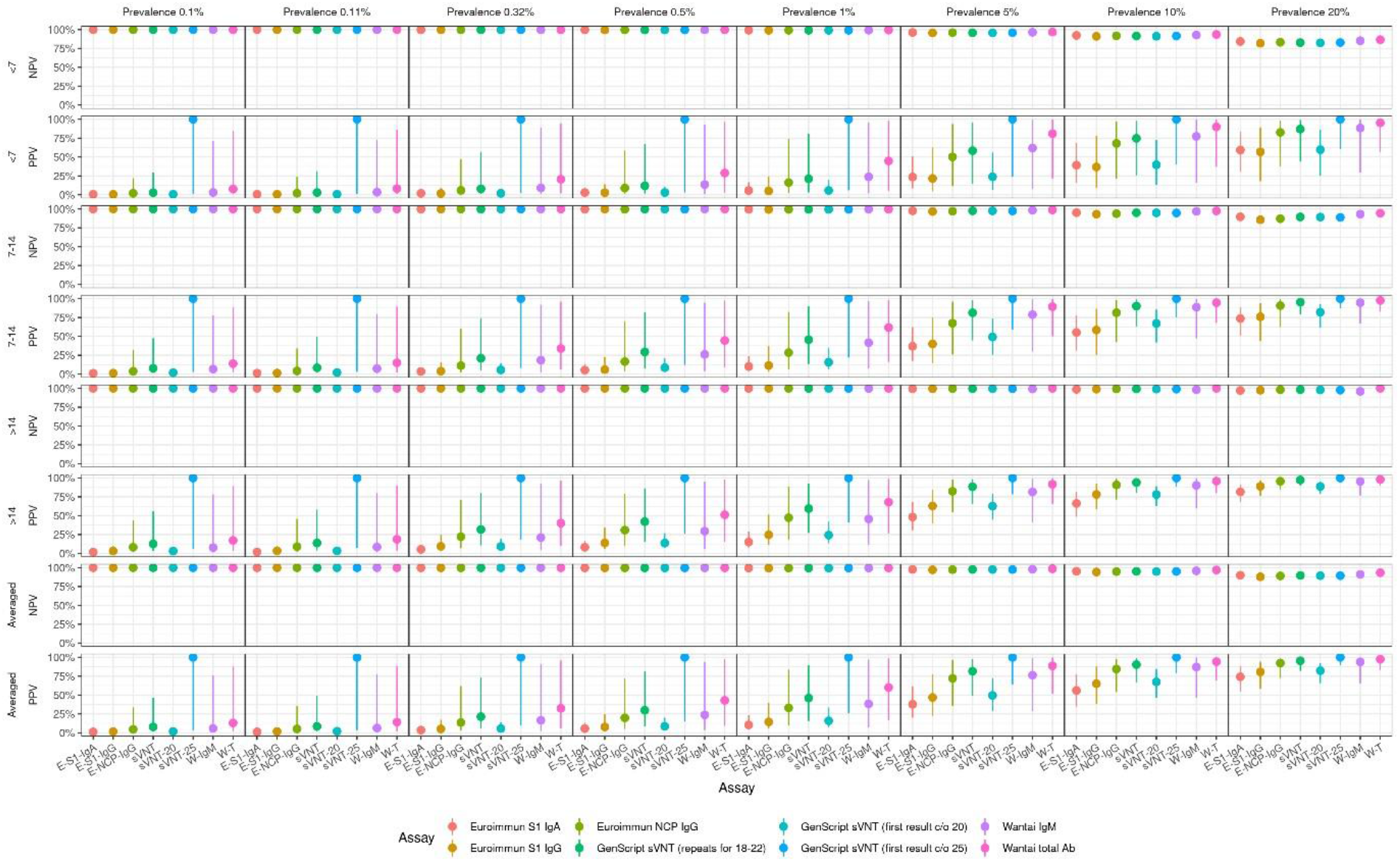
Performance characteristics of the assays across a range of population prevalence estimates: 0.1%, 0.5%, 1%, 5%, 10% and 20%. Abbreviations: E-S1-IgA, Euroimmun S1-IgA; E-S1-IgG, Euroimmun S1-IgG; E-NCP-IgG, Euroimmun NCP-IgG; sVNT, GenScript surrogate virus neutralization test 20% cut-off with repeat testing for equivocal results (18-22); sVNT-20 with 20% cut-off with no repeat testing; sVNT-25 with 25% inhibition cut-off and no repeat testing; W-IgM, Wantai IgM, W-T, Wantai Total Ab.

### Discordance between assays

In the RT-PCR positive group there were 41 discordant results, with discordance increasing with time since disease onset from 8/41 at < 7 days post-onset to 19/41 at > 14 days post-onset. In the population and cross-reactive assessment groups there were 17 and 6 discordant results, respectively. (Supplementary Figure 2– Heat-map).

### Intra and Inter-assay variability

Intra-assay variability was below 10%, ranging from CV=3.1% for the Euroimmun S1-IgA to CV=8.8% for the Euroimmun S1-IgG. Inter-assay variability ranged from Euroimmun NCP-IgG CV=3.1% to Wantai IgM CV=14.9% (Supplementary Table 3a, 3b).

### Assay comparison with microneutralization

Eighty-five samples from RT-PCR positive Panel A were tested by MN (Table 4a). Seventy three percent (62/85) were MN positive. Euroimmun S1-IgA had equal sensitivity with 73% (62/85) samples positive, 67% (57/85) sVNT positive, 67% (56/84) Euroimmun NCP IgG positive and 62% (53/85) Euroimmun S1 IgG positive. Hence MN and Euroimmun S1-IgA had the highest sensitivity. At > 14 days post symptom onset Euroimmun S1 IgA and Euroimmun NCP-IgG had 100% (38/38) sensitivity, followed by MN and Euroimmun S1 IgG at 97% (37/38). sVNT was 95% (36/38) sensitive.

**Table 4a.** Microneutralisation comparison with ELISAs

|  |  | MN | sVNT | Euro<br>S1-IgA | Euro<br>S1-IgG | Euro<br>NCP-IgG |  | MN | Wantai<br>Total<br>Ab | Wantai<br>IgM |
| --- | --- | --- | --- | --- | --- | --- | --- | --- | --- | --- |
| Group |  | Panel A |  |  |  |  |  | Panel B |  |  |
| Sensitivity | No. |  |  |  |  |  |  | No |  |  |
| **Infected-RT-Pos pos/total (%)** | 85 | 62/85<br>(73) | 57/85<br>(67) | 62/85<br>(73) | 53/85<br>(62) | 56/84*<br>(67) | 70 | 55/70<br>(79) | 61/70<br>(87) | 55/70<br>(79) |
| **< 7 days pos/total (%)** | 21 | 9/21<br>(43) | 5/21<br>(24) | 8/21<br>(38) | 4/21<br>(19) | 5/20<br>(25) | 9 | 3/9<br>(33) | 4/9<br>(44) | 3/9<br>(33) |
| **7-14 days pos/total (%)** | 26 | 16/26<br>(62) | 16/26<br>(62) | 16/26<br>(62) | 12/26<br>(46) | 13/26<br>(50) | 20 | 12/20<br>(60) | 16/20<br>(80) | 15/20<br>(75) |
| **> 14 days pos/total (%)** | 38 | 37/38<br>(97) | 36/38<br>(95) | 38/38<br>(100) | 37/38<br>(97) | 38/38<br>(100) | 41 | 40/41<br>(98) | 41/41<br>(100) | 36/41<br>(88) |
| Specificity | No. |  |  |  |  |  |  | No |  |  |
| **Population** | 20 | 0/20 | 0/20 | 11/20 | 2/20 | 0/20 | 14 | 0/14 | 0/14 | 0/14 |
| **Cross-reactive assessment** | 5 | 0/5 | 1/4 | 1/4 | 1/4 | 2/4 | 1 | 0/1 | 1/1 | 1/1 |
| **Total Pos/total (%)** | 25 | 0/25<br>(0) | 1/24<br>(4.2) | 10/24<br>(41.7) | 3/24<br>(12.5) | 2/24<br>(8.3) | 17 | 0/15<br>(0) | 1/15<br>(7.3) | 1/15<br>(7.3) |

**Table 4b.** Microneutralisation comparison with ELISAs (% agreement)

| Group | Euro S1-IgA | Euro S1-IgG | Euro NCP-IgG | sVNT (20% c/o) | svnt-20 (20% c/o/equ) | svnt-25 (25% c/o) | Wantai IgM | Wantai Total Ab |
| --- | --- | --- | --- | --- | --- | --- | --- | --- |
| Sensitivity |  |  |  |  |  |  |  |  |
| Overall % agreement (% best, % worst) |  |  |  |  |  |  |  |  |
| Panel A |  |  |  |  |  |  | Panel B |  |
| Infected |  |  |  |  |  |  |  |  |
| RT-PCR | 80% | 79% | 84% | 84% | 83% | 81% | 91% | 86% |
| Pos | (62%, 91%) | (61%, 92%) | (66%, 95%) | (65%, 95%) | (64%, 94%) | (62%, 94%) | (67%, 99%) | (62%, 97%) |
| Days from symptom onset % agreement (95% CI) [pos/total] |  |  |  |  |  |  |  |  |
|  | 57% | 67% | 75% | 71% | 71% | 71% | 90% | 80% |
| <7 days | (34%, 78%) | (43%, 85%) | (51%, 91%) | (48%, 89%) | (48%, 89%) | (48%, 89%) | (55%,100%) | (44%, 97%) |
|  | [12 / 21] | [14 / 21] | [15 / 20] | [15 / 21] | [15 / 21] | [15 / 21] | [9 / 10] | [8 / 10] |
| 7-14 days | 85% | 77% | 81% | 85% | 85% | 81% | 89% | 79% |
|  | (65%, 96%) | (56%, 91%) | (61%, 93%) | (65%, 96%) | (65%, 96%) | (61%, 93%) | (67%, 99%) | (54%, 94%) |
|  | [22 / 26] | [20 / 26] | [21 / 26] | [22 / 26] | [22 / 26] | [21 / 26] | [17 / 19] | [15 / 19] |
| >14 days | 97% | 95% | 97% | 95% | 92% | 92% | 93% | 98% |
|  | (86%, 100%) | (82%, 99%) | (86%, 100%) | (82%, 99%) | (79%, 98%) | (79%, 98%) | (80%, 98%) | (87%, 100%) |
|  | [37 / 38] | [36 / 38] | [37 / 38] | [36 / 38] | [35 / 38] | [35 / 38] | [38 / 41] | [40 / 41] |
| Specificity |  |  |  |  |  |  |  |  |
| % agreement (95% CI) [pos/total] |  |  |  |  |  |  |  |  |
|  | 55% | 90% | 100% | 100% | 100% | 70% | 100% | 100% |
| Population | (32%, 77%) | (68%, 99%) | (83%,100%) | (83%, 100%) | (83%,100%) | (46%, 88%) | (77%, 100%) | (77%, 100%) |
|  | [11 / 20] | [18 / 20] | [20 / 20] | [20 / 20] | [20 / 20] | [14 / 20] | [14 / 14] | [14 / 14] |
|  | 75% | 80% | 50% | 80% | 80% | 40% | 0% | 0% |
| Cross-reactive | (19%, 99%) | (28%, 99%) | (6.8%, 93%) | (28%, 99%) | (28%, 99%) | (5.3%, 85%) | (0%, 98%) | (0%, 98%) |
|  | [3 / 4] | [4 / 5] | [2 / 4] | [4 / 5] | [4 / 5] | [2 / 5] | [0 / 1] | [0 / 1] |

Group A - 49% of sera (> 14 days post symptom onset); Group B - 69% of sera (>14 days post symptom onset)
Abbreviations: Euro; Euroimmun, sVNT; GenScript surrogate virus neutralization test, CI; Confidence Interval, pos; positive, c/o: cut-off, equ: equivocal.

Seventy samples from RT-PCR positive Panel B were tested by MN. Seventy nine percent (55/70) were MN positive. Wantai Total had highest sensitivity with 87% (61/70) positive. At > 14 days post symptom onset Wantai Total Ab and MN had comparable sensitivities of 100% (41/41) and 98% (40/41) respectively, and Wantai IgM was 88% (36/41) sensitive (Table 4a).

The MN assay specificity for the population and cross-reactive assessment sera groups was 100%.

## DISCUSSION

Reliable COVID-19 serosurveillance is important for guiding the pandemic response [9, 10]. At the individual level, serology can provide a tool for resolving the diagnosis for patients with infections not confirmed by RT-PCR. At the population level, it provides policy-makers with an assessment of overall impact of the pandemic and vaccination efficacy.

Assay sensitivities ranged from 46.7% to 71.6%; and specificities from 95.0% to 99.5% (pre-pandemic population group), and from 83.3% to 97.2% (cross-reactivity assessment group).

Sensitivity for sera collected >14 days post-symptom onset ranged from 83.3% to 100%. Consistent with the biology of an immune response, and as reported by others, when results from all time-points were considered, sensitivity was low, but increased substantially when only sera collected >14 days post-onset was assessed [19], underlining the need to wait a sufficiently long period before confirming infection status by serology. The Euroimmun NCP-IgG and sVNT had good sensitivity, however the highest sensitivity was achieved by Wantai Total Ab at 100%. The Euroimmun NCP-IgG and Wantai IgM had better pre-pandemic population group specificity at 99%, than sVNT. Adjusting the sVNT cut-off to 25% improved its specificity to 100% (Table 2). Wantai Total Ab achieved highest specificity in the cross-reactivity assessment group at 97.2%.

The intra and inter-assay variability of sVNT was not dissimilar to the other assays. All other assays employ an ‘equivocal’ zone, between 10-20% below and above the cut-off. We observed improved sVNT specificity when we varied from the IFU to use a +/-10% repeat equivocal zone.

Cross-reactivity was observed in all assays with sera containing anti-SARS-CoV-1 antibody. Cross reactivity was more commonly seen in the Euroimmun assays, and depending on the assay, in sera positive for MERS, mycoplasma, CMV and dengue antibodies, consistent with other reports [19]. Assays challenged with seasonal coronavirus antibody positive sera were not reactive, and no cross reactivity to sera containing parvovirus B-19 antibodies was seen (Table 3)

Test result discrepancies were observed in the SARS-CoV-2 RT-PCR positive group. In part this appeared attributable to the type of antibody being detected, the antigen targeted in the assay and the assay format. For samples collected > 14 days post-onset, 11 were Wantai IgM negative but Total Ab positive. All 11 samples were collected 21 to > 30 days post onset, suggesting discordance attributable to IgM loss [20]. We retrospectively tested these samples for IgA (results not shown). Six of eleven (55%) were IgA positive, suggesting waning IgM antibody and to a lesser extent IgA. The Wantai Total Ab and sVNT (using a modified population specific cut-off of 25%) demonstrated the highest specificity. This may be because both assays detect Ab to the RBD within the S1-subunit, therefore increasing specificity by exclusion of cross-reacting epitopes outside this domain [19]. sVNT is the only immunoassay correlating with nAb [21,22] although studies have shown samples with Wantai Total Ab positive results with an index ratio > 10 had detectable levels of nAb by VNT [11]. Assay formats which utilise a single detector antibody, for example Wantai Total Ab, may show greater specificity than those using two antibodies in an antigen-antibody-antibody format, such as the Euroimmun assays [23].

With the exception of the Wantai Total Ab, manufacturers for all assays reported higher sensitivities than we demonstrated. However, direct comparison of results is limited by differences in available sample cohorts and sampling timeframes. Our results are consistent with prior reports of sensitivity for Wantai Total Ab of 100% for samples collected > 14 days post-onset [11]; Euroimmun S1-IgG sensitivity post-onset, 43% (7 to 13 days), 67% (14 to 20 days) and 78% (≥ 21 days) and specificity.

96.0% [19]; Euroimmun NCP-IgG 88.89% (>14 days) [22]; and 95.2% (14-17 days), and specificity 94.7% [24]. While other groups have reported slightly higher sVNT sensitivity (95-100%) and specificity 100% [22] than we observed, sensitivity in our assessment was 93.1% (95% CI 84.5-97.7) for sera collected > 14 days post-symptom onset, with specificity 97.1% (pre-pandemic population group). This improved to 100% with use of a modified 25% inhibition cut-off value: and 96.7% for the cross-reactive group.

We compared agreement between the ELISAs and an in-house MN assay, which is the current gold standard to assess protective immunity against SARS-CoV-2 [11]. The Wantai IgM assay had the highest agreement with MN at 91%, followed by Wantai Total Ab at 86% and the lowest was with the Euroimmun S1-IgG at 79%. The sVNT performed according to the IFU gave the best concordance with MN at 84% compared to 83% and 81% for the alternate methods previously described. At > 14 days post symptom onset Wantai Total Ab had the best agreement with MN at 98% followed by Euroimmun S1-IgA, Euro NCP-IgG and sVNT (as per IFU) with (97%, 97% and 95% respectively) and Wantai IgM having the poorest agreement at 93%. MN and ELISA result concordance for sera from the pre-pandemic population group ranged from 55-100% and from 0-80% for the cross-reactive assessment group; MN was the only assay not to show reactivity to the SARS-CoV-1 antibody positive sample.

The sVNT was not as sensitive as MN in our comparison. However, it is simpler to perform than a VNT, does not require a level 3 Biocontainment laboratory, a highly skilled operator nor 5-day test turnaround time, and it has high specificity (if a population adjusted cut-off is employed), and generally good correlation with VNT [25]. Further comparison is required to determine its suitability as an alternative to VNT, in certain settings, particularly as we move into the post-vaccination phase.

In our study, as reported by others, IgA and IgM were detected earlier than IgG, and IgA specificity was lower than IgM or IgG [20]. For clinical testing, the Wantai total antibody assay demonstrated the best overall performance with highest sensitivity, specificity, PPV and NPV. Detecting RBD-Ab, the strong linear correlation between S- and RBD-Ab and ACE-2 receptor binding, suggests nAb is being detected [11, 25]. It is also suitable for population screening where high specificity is required, and high sensitivity is desirable. Other assays such as Euroimmun NCP-IgG have high sensitivity but lower specificity, giving it a low PPV. Further research on the utility of IgA and IgM detection is required, especially as indicators of recent infection. The high specificity sVNT assay was further improved to 100% with cut-off adjustment to 25% for our population. Although sensitivity of the sVNT was lower than the Wantai Total Ab, its specificity makes it a reliable supplemental test to use with screening assays. The assays evaluated here detect antibodies to different viral targets: S, NCP or RBD and emerging humoral response dynamics studies suggest these antibodies have different half-lives [26], an important consideration for serosurveillance. An additional important consideration for assay selection is understanding whether the assay detects non-nAb or nAb (particularly for testing post-vaccination); however further research is required.

The Wantai SARS-CoV-2 total Ab assay was alone among those tested in our hands in achieving sensitivity and specificity very close to that reported by the manufacturer (94.5%; and 100% respectively); one limitation of this assay is the requirement for 100ul of serum versus 10ul for the other assays. In low prevalence populations and with low test PPV’s, serological testing algorithms should use highly sensitive assays for screening such as Wantai Total Ab, followed by supplemental or confirmatory testing with highly specific assays such as sVNT or MN to ensure reliable results. An important component of this study is the comparison of sensitivities of the ELISAs and MN, which should be considered in the context of discordant screening and supplemental results in testing algorithms.

Limitations of our study include the smaller number of convalescent patient RT-PCR-POS confirmed SARS-CoV-2 samples collected > 14 days post-symptom onset used for the Euroimmun and sVNT evaluations; Panel A, 49% (72/147) of samples compared to Panel B, 69% (66/96) of samples used for Wantai Total Ab and IgM assays. A rise in index value/inhibition for all assays was observed over time (Figure 2) however these measurements are semi-quantitative, indicating antibody amount present for comparison between assays, not antibody titre which can only be obtained by sample titration to ‘end-point.’

In conclusion, our study revealed different performance characteristics between six commercially available ELISAs with Wantai Total Ab demonstrating best overall performance. As we move into the post-vaccination phase consideration should be given to the use of GenScript sVNT in certain settings, as an alternative to VNT. While a positive ELISA result with neutralising activity, even if specific, provides evidence of prior SARS-CoV-2 infection, more research is required to determine correlates of protection, antibody longevity and the role played by the adaptive immune system.

## Data Availability

All data regarding the study is available on request.

## ACKNOWLEDGEMENTS

The authors thank the Serology Laboratory at VIDRL, particularly Di Karamalakis, The Viral Identification Laboratory and the Information Technology department, particularly Dallas Wilson. We thank Dr. Allen Cheng, Dr. Maryza Graham and Megan Wieringa from Monash Medical Centre and Dr. Katherine Kedzierska from The Department of Microbiology and Immunology, The University of Melbourne at The Doherty Institute, for referring samples to VIDRL for COVID-19 investigation. Annette Fox from WHO Collaborating Centre for Reference and Research on Influenza, VIDRL, Doherty Institute for her contribution to the study. Prof. Lin Fa Wang for supplying us with the sVNT assay for evaluation.

The Victorian Infectious Diseases Reference Laboratory is supported by the Victorian Department of Health and Human Services. The WHO Collaborating Centre for Reference and Research on Influenza is supported by the Australian Government Department of Health.

## Conflicts of interest

The authors declare no competing interests.

## Notes

### Competing Interest Statement

The authors have declared no competing interest.

### Funding Statement

No external funding was received.

### Author Declarations

Project ethical approval for RMH specimens was obtained from Melbourne Health Human Research Ethics Committee (RMH HREC QA2020052).

