## Supplementary Data for "Evaluation of six commercial SARS-CoV-2 Enzyme-Linked Immunosorbent assays for clinical testing and serosurveillance"

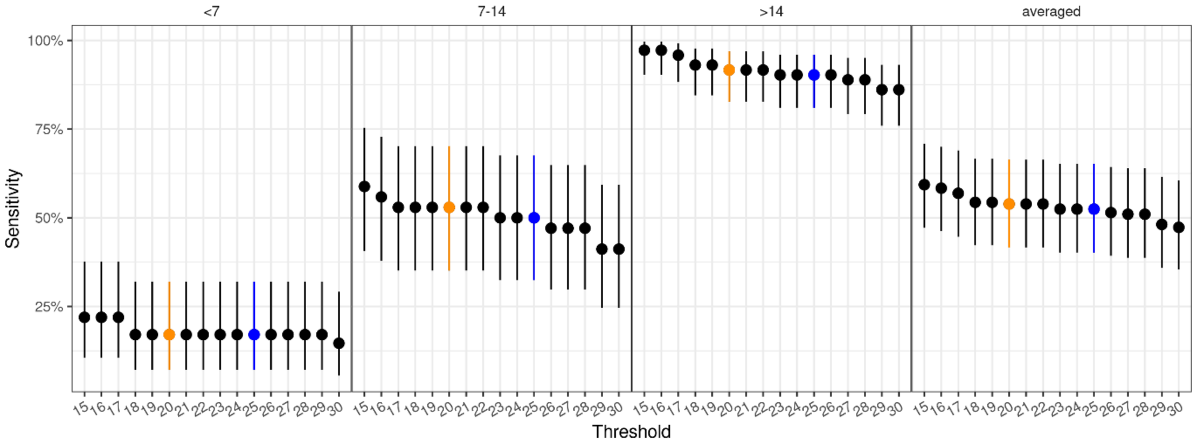


**Figure 1a – GenScript sVNT sensitivity analysis at different cut-off thresholds post symptom onset: < 7days, 7-14 days, > 14 days and averaged.**

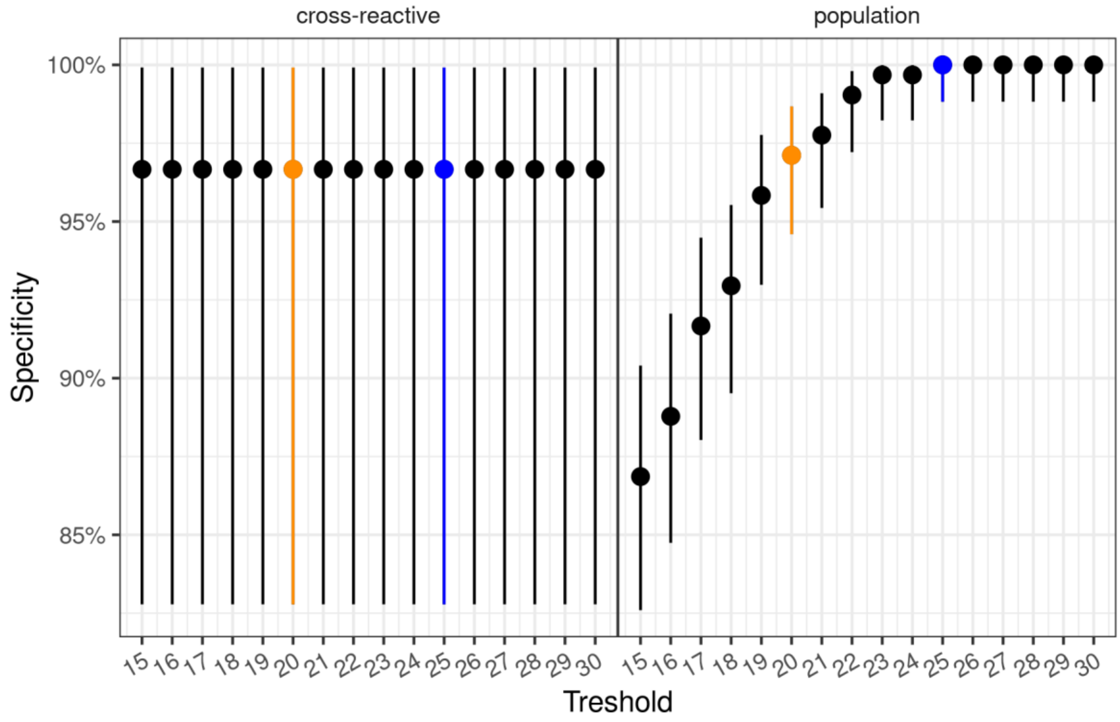


**Figure 1b – GenScript sVNT specificity analysis at different cut-off thresholds for population cohort and cross-reactive assessment cohort.**

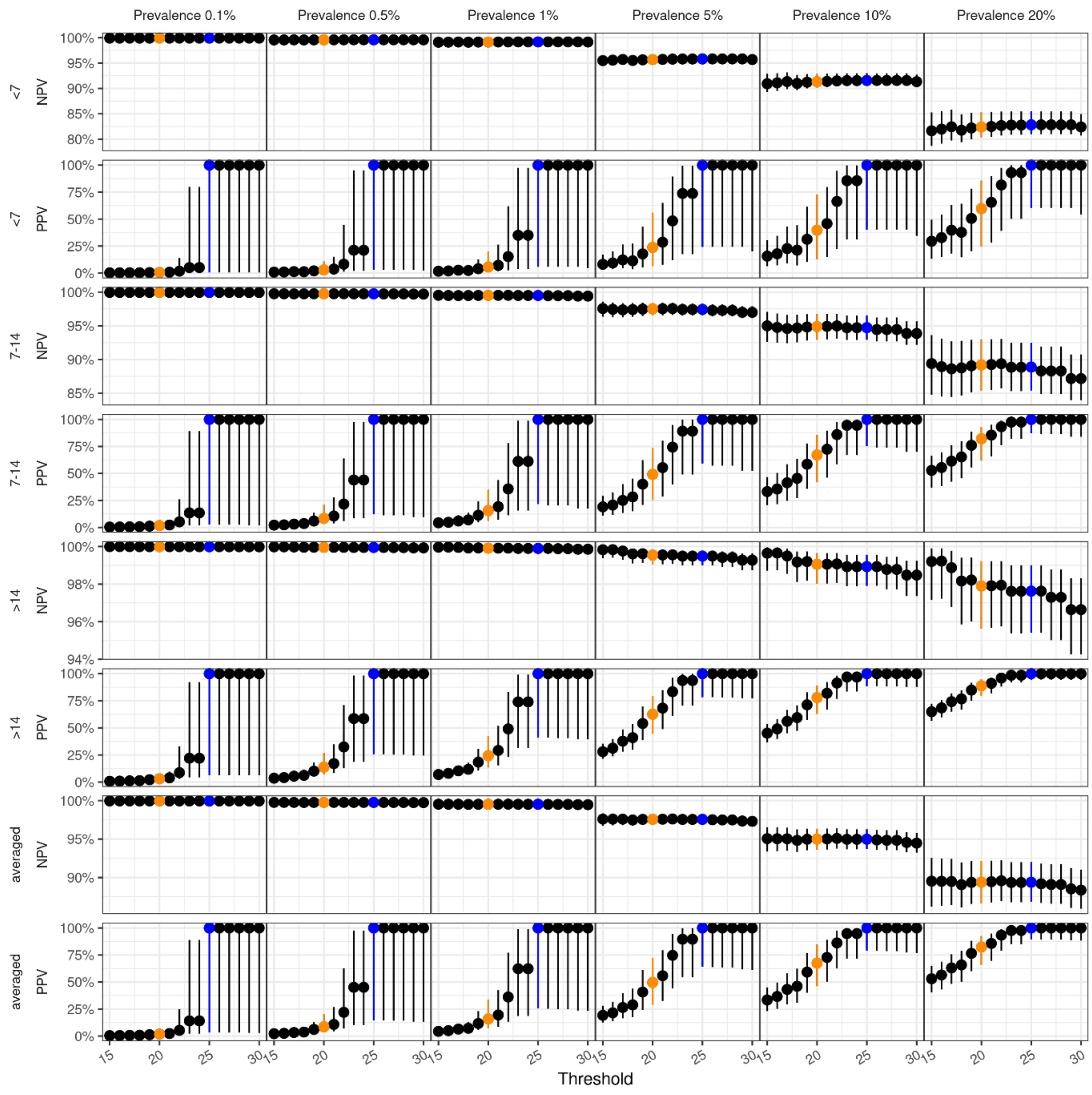


**Figure 1c – GenScript sVNT Positive PPV and NPV at different population prevalence rates: 0.1%, 0.5%, 1%, 5%, 10% and 20%.**

**Table 1a – GenScript sVNT sensitivity data analysis for cut-off adjustment**

| Sensitivity [95% CI] | < 7 days | 7 – 14 days | >14 days | Averaged |
|---|---|---|---|---|
| **Cut-off (%)** | | | | |
| **15** | 22% [10.6% - 37.6%] (9 / 41) | 58.8% [40.7% - 75.4%] (20 / 34) | 97.2% [90.3% - 99.7%] (70 / 72) | 59.3% [47.2% - 70.9%] |
| **16** | 22% [10.6% - 37.6%] (9 / 41) | 55.9% [37.9% - 72.8%] (19 / 34) | 97.2% [90.3% - 99.7%] (70 / 72) | 58.4% [46.3% - 70%] |
| **17** | 22% [10.6% - 37.6%] (9 / 41) | 52.9% [35.1% - 70.2%] (18 / 34) | 95.8% [88.3% - 99.1%] (69 / 72) | 56.9% [44.7% - 69%] |
| **18** | 17.1% [7.2% - 32.1%] (7 / 41) | 52.9% [35.1% - 70.2%] (18 / 34) | 93.1% [84.5% - 97.7%] (67 / 72) | 54.4% [42.3% - 66.7%] |
| **19** | 17.1% [7.2% - 32.1%] (7 / 41) | 52.9% [35.1% - 70.2%] (18 / 34) | 93.1% [84.5% - 97.7%] (67 / 72) | 54.4% [42.3% - 66.7%] |
| **20** | 17.1% [7.2% - 32.1%] (7 / 41) | 52.9% [35.1% - 70.2%] (18 / 34) | 91.7% [82.7% - 96.9%] (66 / 72) | 53.9% [41.7% - 66.4%] |
| **21** | 17.1% [7.2% - 32.1%] (7 / 41) | 52.9% [35.1% - 70.2%] (18 / 34) | 91.7% [82.7% - 96.9%] (66 / 72) | 53.9% [41.7% - 66.4%] |
| **22** | 17.1% [7.2% - 32.1%] (7 / 41) | 52.9% [35.1% - 70.2%] (18 / 34) | 91.7% [82.7% - 96.9%] (66 / 72) | 53.9% [41.7% - 66.4%] |
| **23** | 17.1% [7.2% - 32.1%] (7 / 41) | 50% [32.4% - 67.6%] (17 / 34) | 90.3% [81% - 96%] (65 / 72) | 52.5% [40.2% - 65.2%] |
| **24** | 17.1% [7.2% - 32.1%] (7 / 41) | 50% [32.4% - 67.6%] (17 / 34) | 90.3% [81% - 96%] (65 / 72) | 52.5% [40.2% - 65.2%] |
| **25** | 17.1% [7.2% - 32.1%] (7 / 41) | 50% [32.4% - 67.6%] (17 / 34) | 90.3% [81% - 96%] (65 / 72) | 52.5% [40.2% - 65.2%] |
| **26** | 17.1% [7.2% - 32.1%] (7 / 41) | 47.1% [29.8% - 64.9%] (16 / 34) | 90.3% [81% - 96%] (65 / 72) | 51.5% [39.3% - 64.3%] |
| **27** | 17.1% [7.2% - 32.1%] (7 / 41) | 47.1% [29.8% - 64.9%] (16 / 34) | 88.9% [79.3% - 95.1%] (64 / 72) | 51% [38.7% - 64%] |
| **28** | 17.1% [7.2% - 32.1%] (7 / 41) | 47.1% [29.8% - 64.9%] (16 / 34) | 88.9% [79.3% - 95.1%] (64 / 72) | 51% [38.7% - 64%] |
| **29** | 17.1% [7.2% - 32.1%] (7 / 41) | 41.2% [24.6% - 59.3%] (14 / 34) | 86.1% [75.9% - 93.1%] (62 / 72) | 48.1% [35.9% - 61.5%] |
| **30** | 14.6% [5.6% - 29.2%] (6 / 41) | 41.2% [24.6% - 59.3%] (14 / 34) | 86.1% [75.9% - 93.1%] (62 / 72) | 47.3% [35.4% - 60.5%] |

**Table 1b – GenScript sVNT specificity data analysis for cut-off adjustment**

| Specificity [95% CI] | Cross-reactive cohort | Population cohort |
|---|---|---|
| **Cut-off (%)** | | |
| **15** | 96.7% [82.8% - 99.9%] (29 / 30) | 86.9% [82.6% - 90.4%] (271 / 312) |
| **16** | 96.7% [82.8% - 99.9%] (29 / 30) | 88.8% [84.7% - 92.1%] (277 / 312) |
| **17** | 96.7% [82.8% - 99.9%] (29 / 30) | 91.7% [88% - 94.5%] (286 / 312) |
| **18** | 96.7% [82.8% - 99.9%] (29 / 30) | 92.9% [89.5% - 95.5%] (290 / 312) |
| **19** | 96.7% [82.8% - 99.9%] (29 / 30) | 95.8% [93% - 97.8%] (299 / 312) |
| **20** | 96.7% [82.8% - 99.9%] (29 / 30) | 97.1% [94.6% - 98.7%] (303 / 312) |
| **21** | 96.7% [82.8% - 99.9%] (29 / 30) | 97.8% [95.4% - 99.1%] (305 / 312) |
| **22** | 96.7% [82.8% - 99.9%] (29 / 30) | 99% [97.2% - 99.8%] (309 / 312) |
| **23** | 96.7% [82.8% - 99.9%] (29 / 30) | 99.7% [98.2% - 100%] (311 / 312) |
| **24** | 96.7% [82.8% - 99.9%] (29 / 30) | 99.7% [98.2% - 100%] (311 / 312) |
| **25** | 96.7% [82.8% - 99.9%] (29 / 30) | 100% [98.8% - 100%] (312 / 312) |
| **26** | 96.7% [82.8% - 99.9%] (29 / 30) | 100% [98.8% - 100%] (312 / 312) |
| **27** | 96.7% [82.8% - 99.9%] (29 / 30) | 100% [98.8% - 100%] (312 / 312) |
| **28** | 96.7% [82.8% - 99.9%] (29 / 30) | 100% [98.8% - 100%] (312 / 312) |
| **29** | 96.7% [82.8% - 99.9%] (29 / 30) | 100% [98.8% - 100%] (312 / 312) |
| **30** | 96.7% [82.8% - 99.9%] (29 / 30) | 100% [98.8% - 100%] (312 / 312) |

**Supplementary Table 2 – Positive Predictive & Negative Predictive Values for assays across a range of population prevalence estimates: 0.1%, 0.5%, 1%, 5%, 10% and 20% and days post symptom onset.**

| Symptom onset | char | Preva-lence | Euro S1-IgA | Euro S1-IgG | Euro NCP-IgG | sVNT | sVNT - 20 | sVNT - 25 | Wantai IgM | Wantai Total Ab |
|---|---|---|---|---|---|---|---|---|---|---|
| | | | % estimate [95% CI] | | | | | | | |
| <7 | ppv | 0.001 | 0.6% [0.2% - 1.9%] | 0.5% [0.1% - 3.1%] | 1.9% [0.2% - 21.9%] | 2.6% [0.3% - 29.2%] | 0.6% [0.1% - 2.4%] | 100% [0.6% - 100%] | 3% [0.2% - 70.8%] | 7.4% [0.5% - 85%] |
| <7 | npv | 0.001 | 99.9% [99.9% - 99.9%] | 99.9% [99.9% - 99.9%] | 99.9% [99.9% - 99.9%] | 99.9% [99.9% - 99.9%] | 99.9% [99.9% - 99.9%] | 99.9% [99.9% - 99.9%] | 99.9% [99.9% - 100%] | 99.9% [99.9% - 100%] |
| <7 | ppv | 0.005 | 2.8% [0.9% - 9%] | 2.6% [0.4% - 13.8%] | 8.8% [1.2% - 58.5%] | 11.8% [1.5% - 67.5%] | 2.9% [0.7% - 10.8%] | 100% [3% - 100%] | 13.4% [0.8% - 92.4%] | 28.8% [2.6% - 96.6%] |
| <7 | npv | 0.005 | 99.6% [99.5% - 99.7%] | 99.6% [99.5% - 99.6%] | 99.6% [99.5% - 99.7%] | 99.6% [99.5% - 99.7%] | 99.6% [99.5% - 99.7%] | 99.6% [99.5% - 99.7%] | 99.6% [99.5% - 99.8%] | 99.7% [99.6% - 99.8%] |
| <7 | ppv | 0.01 | 5.6% [1.7% - 16.5%] | 5% [0.9% - 24.4%] | 16.2% [2.4% - 73.9%] | 21.2% [3% - 80.6%] | 5.6% [1.3% - 19.6%] | 100% [5.8% - 100%] | 23.7% [1.7% - 96.1%] | 44.8% [5% - 98.3%] |
| <7 | npv | 0.01 | 99.3% [99.1% - 99.4%] | 99.1% [99% - 99.3%] | 99.2% [99.1% - 99.4%] | 99.2% [99% - 99.3%] | 99.1% [99% - 99.3%] | 99.2% [99.1% - 99.3%] | 99.3% [99% - 99.6%] | 99.4% [99.1% - 99.7%] |
| <7 | ppv | 0.05 | 23.5% [8.3% - 50.8%] | 21.6% [4.4% - 62.7%] | 50.1% [11.3% - 93.7%] | 58.4% [14.1% - 95.6%] | 23.8% [6.5% - 56%] | 100% [24.3% - 100%] | 61.8% [8.1% - 99.2%] | 80.9% [21.7% - 99.7%] |
| <7 | npv | 0.05 | 96.2% [95.4% - 97.1%] | 95.6% [95% - 96.4%] | 95.9% [95.3% - 96.7%] | 95.8% [95.2% - 96.5%] | 95.7% [95.1% - 96.5%] | 95.8% [95.3% - 96.5%] | 96.5% [95.2% - 98%] | 96.8% [95.6% - 98.4%] |
| <7 | ppv | 0.1 | 39.3% [16.1% - 68.5%] | 36.8% [8.8% - 78%] | 68% [21.2% - 96.9%] | 74.7% [25.7% - 97.9%] | 39.7% [12.8% - 72.9%] | 100% [40.3% - 100%] | 77.4% [15.6% - 99.6%] | 89.9% [36.9% - 99.8%] |
| <7 | npv | 0.1 | 92.4% [90.7% - 94.2%] | 91.1% [89.9% - 92.6%] | 91.8% [90.5% - 93.3%] | 91.5% [90.4% - 93%] | 91.3% [90.2% - 92.9%] | 91.6% [90.5% - 93%] | 92.8% [90.3% - 95.9%] | 93.6% [91% - 96.6%] |
| <7 | ppv | 0.2 | 59.3% [30.2% - 83%] | 56.7% [17.9% - 88.9%] | 82.7% [37.8% - 98.6%] | 86.9% [43.8% - 99%] | 59.7% [24.9% - 85.8%] | 100% [60.3% - 100%] | 88.5% [29.4% - 99.8%] | 95.3% [56.8% - 99.9%] |
| <7 | npv | 0.2 | 84.3% [81.2% - 87.8%] | 82% [79.9% - 84.8%] | 83.2% [80.9% - 86.1%] | 82.7% [80.8% - 85.5%] | 82.4% [80.3% - 85.3%] | 82.8% [81% - 85.5%] | 85.1% [80.6% - 91.2%] | 86.6% [81.9% - 92.7%] |
| 7-14 | ppv | 0.001 | 1.1% [0.4% - 3%] | 1.2% [0.3% - 5.5%] | 3.8% [0.7% - 31.8%] | 7.6% [1.5% - 47.5%] | 1.8% [0.6% - 5%] | 100% [2.7% - 100%] | 6.6% [0.8% - 78%] | 13.8% [1.9% - 88.5%] |
| 7-14 | npv | 0.001 | 100% [99.9% - 100%] | 99.9% [99.9% - 100%] | 99.9% [99.9% - 100%] | 100% [99.9% - 100%] | 100% [99.9% - 100%] | 99.9% [99.9% - 100%] | 100% [99.9% - 100%] | 100% [99.9% - 100%] |
| 7-14 | ppv | 0.005 | 5.3% [2% - 13.6%] | 6% [1.5% - 22.8%] | 16.5% [3.2% - 70.1%] | 29.3% [7.1% - 81.9%] | 8.4% [3.2% - 21%] | 100% [12.2% - 100%] | 26.2% [3.9% - 94.7%] | 44.5% [8.7% - 97.5%] |
| 7-14 | npv | 0.005 | 99.8% [99.7% - 99.9%] | 99.7% [99.6% - 99.8%] | 99.7% [99.6% - 99.8%] | 99.8% [99.7% - 99.9%] | 99.8% [99.7% - 99.8%] | 99.7% [99.7% - 99.8%] | 99.9% [99.7% - 99.9%] | 99.9% [99.7% - 100%] |
| 7-14 | ppv | 0.01 | 10.1% [3.9% - 24%] | 11.3% [3% - 37.2%] | 28.4% [6.3% - 82.5%] | 45.5% [13.4% - 90.1%] | 15.6% [6.2% - 34.8%] | 100% [21.8% - 100%] | 41.6% [7.6% - 97.3%] | 61.7% [16.1% - 98.7%] |
| 7-14 | npv | 0.01 | 99.5% [99.3% - 99.7%] | 99.3% [99.1% - 99.5%] | 99.4% [99.2% - 99.6%] | 99.5% [99.3% - 99.7%] | 99.5% [99.3% - 99.7%] | 99.5% [99.3% - 99.7%] | 99.7% [99.4% - 99.9%] | 99.8% [99.5% - 99.9%] |

| Symptom onset | char | Preva-lence | Euro S1-IgA | Euro S1-IgG | Euro NCP-IgG | sVNT | sVNT - 20 | sVNT - 25 | Wantai IgM | Wantai Total Ab |
|---|---|---|---|---|---|---|---|---|---|---|
| | | | | | | % estimate [95% CI] | | | | |
| 7-14 | ppv | 0.05 | 36.9% [17.6% - 62.2%] | 39.9% [14% - 75.5%] | 67.4% [25.8% - 96.1%] | 81.3% [44.6% - 97.9%] | 49.1% [25.5% - 73.6%] | 100% [59.2% - 100%] | 78.8% [29.9% - 99.5%] | 89.4% [50% - 99.8%] |
| 7-14 | npv | 0.05 | 97.6% [96.5% - 98.6%] | 96.6% [95.7% - 97.6%] | 97% [96% - 97.9%] | 97.6% [96.6% - 98.5%] | 97.5% [96.5% - 98.4%] | 97.4% [96.5% - 98.3%] | 98.5% [97% - 99.5%] | 98.8% [97.4% - 99.6%] |
| 7-14 | ppv | 0.1 | 55.3% [31.1% - 77.7%] | 58.4% [25.5% - 86.7%] | 81.4% [42.3% - 98.1%] | 90.2% [63% - 99%] | 67.1% [41.9% - 85.5%] | 100% [75.4% - 100%] | 88.7% [47.3% - 99.7%] | 94.7% [67.9% - 99.9%] |
| 7-14 | npv | 0.1 | 95.1% [92.9% - 97%] | 93.1% [91.3% - 95%] | 93.8% [92% - 95.7%] | 95% [93.1% - 96.8%] | 94.9% [92.9% - 96.8%] | 94.7% [92.9% - 96.5%] | 96.8% [93.8% - 98.9%] | 97.4% [94.6% - 99.2%] |
| 7-14 | ppv | 0.2 | 73.5% [50.4% - 88.7%] | 76% [43.6% - 93.6%] | 90.8% [62.3% - 99.2%] | 95.4% [79.3% - 99.6%] | 82.1% [61.9% - 93%] | 100% [87.3% - 100%] | 94.6% [66.9% - 99.9%] | 97.6% [82.6% - 99.9%] |
| 7-14 | npv | 0.2 | 89.6% [85.4% - 93.5%] | 85.7% [82.3% - 89.5%] | 87.1% [83.6% - 90.8%] | 89.4% [85.8% - 93.1%] | 89.2% [85.4% - 93%] | 88.9% [85.4% - 92.5%] | 93.1% [87.1% - 97.5%] | 94.4% [88.6% - 98.3%] |
| >14 | ppv | 0.001 | 1.7% [0.8% - 3.9%] | 3.1% [1.3% - 9.5%] | 8.2% [2.2% - 43.5%] | 12.7% [3.6% - 55.7%] | 3.1% [1.5% - 6.8%] | 100% [6.5% - 100%] | 7.7% [1.3% - 78.3%] | 17.3% [3.5% - 89.2%] |
| >14 | npv | 0.001 | 100% [100% - 100%] | 100% [100% - 100%] | 100% [100% - 100%] | 100% [100% - 100%] | 100% [100% - 100%] | 100% [100% - 100%] | 100% [100% - 100%] | 100% [100% - 100%] |
| >14 | ppv | 0.005 | 8.2% [4.1% - 17%] | 14% [6% - 34.6%] | 30.9% [10.2% - 79.4%] | 42.2% [15.6% - 86.3%] | 13.8% [7.1% - 26.8%] | 100% [25.7% - 100%] | 29.5% [6.2% - 94.8%] | 51.2% [15.3% - 97.6%] |
| >14 | npv | 0.005 | 99.9% [99.9% - 100%] | 99.9% [99.9% - 100%] | 100% [99.9% - 100%] | 100% [99.9% - 100%] | 100% [99.9% - 100%] | 100% [99.9% - 100%] | 99.9% [99.9% - 100%] | 100% [100% - 100%] |
| >14 | ppv | 0.01 | 15.2% [7.9% - 29.2%] | 24.6% [11.3% - 51.5%] | 47.3% [18.6% - 88.6%] | 59.5% [27.1% - 92.7%] | 24.3% [13.4% - 42.4%] | 100% [41% - 100%] | 45.7% [11.8% - 97.3%] | 67.9% [26.6% - 98.8%] |
| >14 | npv | 0.01 | 99.9% [99.8% - 99.9%] | 99.9% [99.8% - 100%] | 99.9% [99.8% - 100%] | 99.9% [99.8% - 100%] | 99.9% [99.8% - 100%] | 99.9% [99.8% - 100%] | 99.8% [99.7% - 99.9%] | 100% [99.9% - 100%] |
| >14 | ppv | 0.05 | 48.2% [30.9% - 68.3%] | 63% [40% - 84.7%] | 82.4% [54.4% - 97.6%] | 88.4% [66% - 98.5%] | 62.6% [44.6% - 79.3%] | 100% [78.4% - 100%] | 81.4% [41.1% - 99.5%] | 91.7% [65.4% - 99.8%] |
| >14 | npv | 0.05 | 99.4% [98.8% - 99.7%] | 99.5% [98.9% - 99.8%] | 99.6% [99.2% - 99.9%] | 99.6% [99.2% - 99.9%] | 99.6% [99% - 99.8%] | 99.5% [99% - 99.8%] | 99.1% [98.5% - 99.5%] | 100% [99.7% - 100%] |
| >14 | ppv | 0.1 | 66.3% [48.6% - 82%] | 78.2% [58.4% - 92.1%] | 90.8% [71.6% - 98.8%] | 94.2% [80.4% - 99.3%] | 77.9% [63% - 89%] | 100% [88.4% - 100%] | 90.3% [59.5% - 99.8%] | 95.9% [79.9% - 99.9%] |
| >14 | npv | 0.1 | 98.7% [97.5% - 99.4%] | 98.9% [97.8% - 99.6%] | 99.2% [98.2% - 99.7%] | 99.2% [98.3% - 99.7%] | 99.1% [98% - 99.6%] | 98.9% [97.9% - 99.6%] | 98.2% [96.8% - 99.1%] | 100% [99.4% - 100%] |
| >14 | ppv | 0.2 | 81.5% [68% - 91.1%] | 89% [76% - 96.3%] | 95.7% [85% - 99.5%] | 97.3% [90.2% - 99.7%] | 88.8% [79.3% - 94.8%] | 100% [94.5% - 100%] | 95.4% [76.8% - 99.9%] | 98.1% [90% - 100%] |
| >14 | npv | 0.2 | 97.2% [94.6% - 98.8%] | 97.6% [95.2% - 99%] | 98.3% [96.1% - 99.4%] | 98.3% [96.2% - 99.4%] | 97.9% [95.6% - 99.2%] | 97.6% [95.4% - 99%] | 96% [93.1% - 97.9%] | 100% [98.6% - 100%] |
| averaged | ppv | 0.001 | 1.1% [0.5% - 3%] | 1.6% [0.6% - 6.1%] | 4.7% [1% - 33.6%] | 7.8% [1.8% - 46.2%] | 1.8% [0.8% - 4.8%] | 100% [3.3% - 100%] | 5.8% [0.8% - 76.2%] | 13% [2% - 87.8%] |
| averaged | npv | 0.001 | 100% [99.9% - 100%] | 99.9% [99.9% - 100%] | 100% [99.9% - 100%] | 100% [99.9% - 100%] | 100% [99.9% - 100%] | 100% [99.9% - 100%] | 100% [99.9% - 100%] | 100% [100% - 100%] |

| Symptom onset | char | Preva-lence | Euro S1-IgA | Euro S1-IgG | Euro NCP-IgG | sVNT | sVNT - 20 | sVNT - 25 | Wantai IgM | Wantai Total Ab |
|---|---|---|---|---|---|---|---|---|---|---|
| | | | | | | % estimate [95% CI] | | | | |
| averaged | ppv | 0.005 | 5.5% [2.3% - 13.3%] | 7.8% [2.7% - 24.7%] | 19.8% [5% - 71.7%] | 29.9% [8.5% - 81.2%] | 8.6% [3.7% - 20.1%] | 100% [14.7% - 100%] | 23.6% [3.7% - 94.1%] | 42.9% [9.1% - 97.3%] |
| averaged | npv | 0.005 | 99.8% [99.7% - 99.9%] | 99.7% [99.7% - 99.8%] | 99.8% [99.7% - 99.8%] | 99.8% [99.7% - 99.8%] | 99.8% [99.7% - 99.8%] | 99.8% [99.7% - 99.8%] | 99.8% [99.7% - 99.9%] | 99.9% [99.8% - 99.9%] |
| averaged | ppv | 0.01 | 10.4% [4.6% - 23.6%] | 14.5% [5.3% - 39.7%] | 33.2% [9.6% - 83.6%] | 46.1% [15.7% - 89.7%] | 15.9% [7.2% - 33.6%] | 100% [25.7% - 100%] | 38.3% [7.2% - 97%] | 60.2% [16.8% - 98.6%] |
| averaged | npv | 0.01 | 99.6% [99.4% - 99.7%] | 99.4% [99.3% - 99.6%] | 99.5% [99.4% - 99.6%] | 99.5% [99.4% - 99.7%] | 99.5% [99.4% - 99.7%] | 99.5% [99.4% - 99.6%] | 99.6% [99.4% - 99.8%] | 99.7% [99.5% - 99.9%] |
| averaged | ppv | 0.05 | 37.8% [20% - 61.7%] | 46.8% [22.6% - 77.4%] | 72.1% [35.7% - 96.4%] | 81.7% [49.2% - 97.8%] | 49.6% [28.9% - 72.5%] | 100% [64.3% - 100%] | 76.4% [28.8% - 99.4%] | 88.7% [51.3% - 99.7%] |
| averaged | npv | 0.05 | 97.7% [96.9% - 98.5%] | 97.2% [96.5% - 97.9%] | 97.5% [96.8% - 98.1%] | 97.6% [97% - 98.3%] | 97.6% [96.9% - 98.2%] | 97.6% [96.9% - 98.2%] | 98% [96.9% - 99%] | 98.5% [97.5% - 99.3%] |
| averaged | ppv | 0.1 | 56.2% [34.6% - 77.3%] | 65% [38.1% - 87.9%] | 84.5% [54% - 98.3%] | 90.4% [67.2% - 99%] | 67.5% [46.1% - 84.7%] | 100% [79.2% - 100%] | 87.2% [46% - 99.7%] | 94.3% [69% - 99.9%] |
| averaged | npv | 0.1 | 95.3% [93.6% - 96.8%] | 94.3% [92.9% - 95.7%] | 94.8% [93.5% - 96.2%] | 95.1% [93.8% - 96.4%] | 95% [93.6% - 96.4%] | 95% [93.7% - 96.3%] | 95.9% [93.6% - 97.9%] | 96.9% [94.9% - 98.6%] |
| averaged | ppv | 0.2 | 74.3% [54.3% - 88.4%] | 80.7% [58.1% - 94.2%] | 92.5% [72.5% - 99.2%] | 95.5% [82.1% - 99.5%] | 82.4% [65.8% - 92.6%] | 100% [89.5% - 100%] | 93.9% [65.7% - 99.9%] | 97.4% [83.4% - 99.9%] |
| averaged | npv | 0.2 | 90% [86.7% - 93.1%] | 88% [85.3% - 90.7%] | 89.1% [86.4% - 91.8%] | 89.7% [87.1% - 92.3%] | 89.4% [86.6% - 92.2%] | 89.4% [86.9% - 92%] | 91.2% [86.7% - 95.4%] | 93.4% [89.2% - 96.9%] |

Performance characteristics of the assays across a range of population prevalence estimates: 0.1%, 0.5%, 1%, 5%, 10% and 20%. PPV; Positive Predictive Value. NPV; Negative Predictive Value Abbreviations: Euro; Euroimmun, sVNT, GenScript surrogate virus neutralization test 20% cut-off with repeat testing for equivocal results (18-22); svnt-20 with 20% cut-off with no repeat testing; svnt-25 with 25% inhibition cut-off and no repeat testing.

The code can be found at

https://github.com/khvorov45/roc

**Supplementary Table 3(a). Intra-assay variability**

| Assay | Mean Index (S/CO) [95% CI] | Standard Deviation | Coefficient of Variation (%) |
|---|---|---|---|
| Euro S1-IgA | 2.80 [2.75-2.90] | 0.1 | 3.1 |
| Euro S1-IgG | 2.20 [1.93-2.26] | 0.2 | 8.8 |
| Euro NCP-IgG | 3.10 [3.00-3.20] | 0.1 | 4.1 |
| GenScript sVNT | 0.50 [0.50-0.53] | 0.0 | 5.6 |
| Wantai Total Ab | 2.10 [2.04-2.14] | 0.1 | 5.0 |
| Wantai IgM | 1.60 [1.56-1.69] | 0.1 | 6.7 |

**Supplementary Table 3(b). Inter-assay variability**

| Assay | Mean Index (S/CO) [95% CI] | Standard Deviation | Coefficient of Variation (%) |
|---|---|---|---|
| Euro S1-IgA | 1.90 [1.77-2.05] | 0.2 | 9.2 |
| Euro S1-IgG | 2.68 [2.44-2.92] | 0.3 | 10.4 |
| Euro NCP-IgG | 2.80 [2.70-2.90] | 0.1 | 3.1 |
| GenScript sVNT | 0.47 [0.45-0.50] | 0.0 | 10.4 |
| Wantai Total Ab | 4.79 [4.54-5.03] | 0.3 | 5.9 |
| Wantai IgM | 4.2 [4.20-5.50] | 0.7 | 14.9 |

Euro; Euroimmun, sVNT; GenScript surrogate virus neutralization test

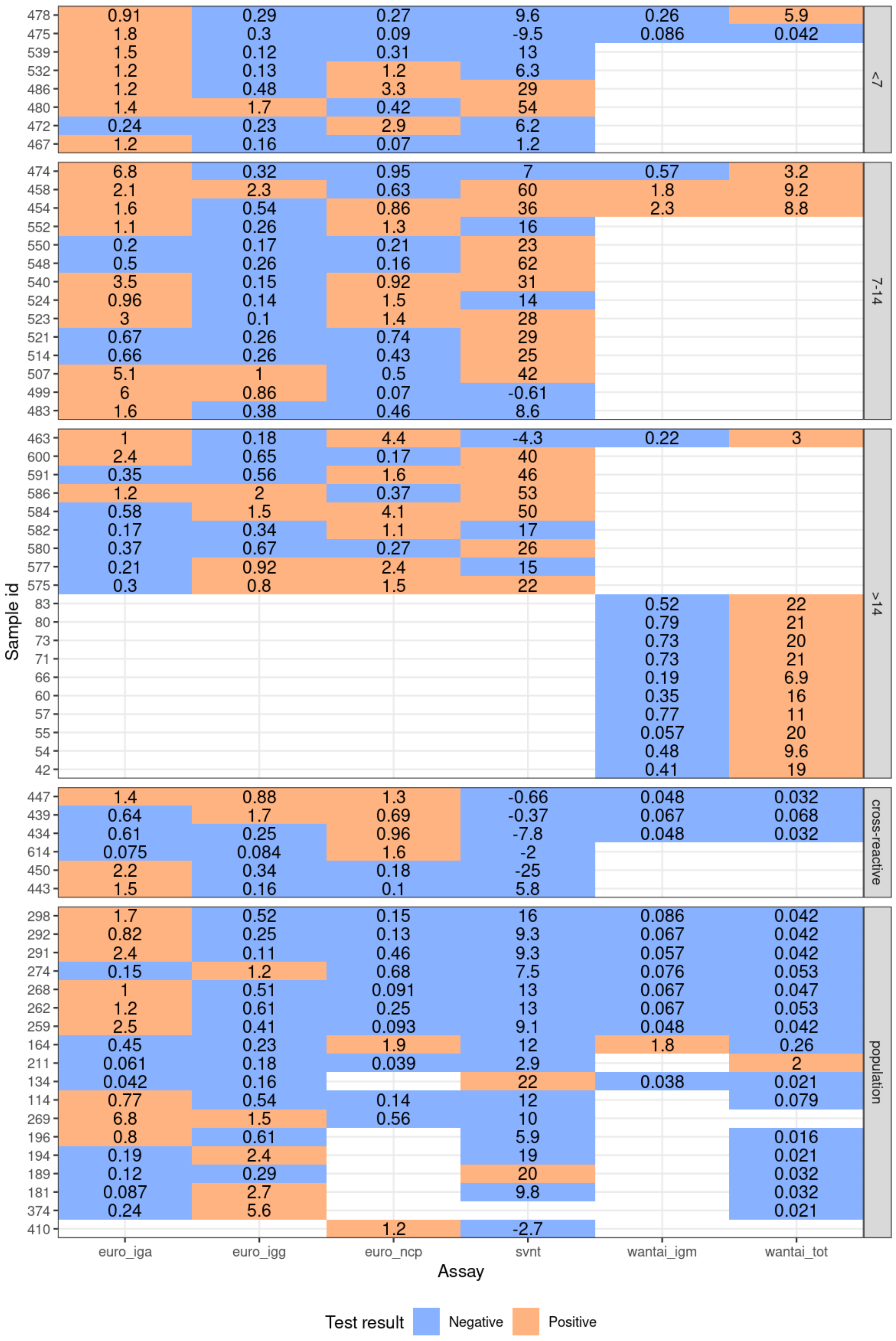


**Supplementary Figure 2** – Heatmap of discordant results with numbers corresponding to the measurement. Measure is index value (sample optical density (OD) value/ cut-off OD) for all assays except sVNT. sVNT measure is %inhibition, calculated as per IFU. euro_iga, Euroimmun S1-IgA; euro_igg, Euroimmun S1-IgG; euro_ncp, Euroimmun NCP-IgG; svnt, GenScript surrogate virus neutralization test; wantai_igm, wantai IgM, wantai_tot, Wantai Total Ab.
